# Higher distal proximal skin temperature is associated with reduced bedtime vigilance in young people with major depressive disorder

**DOI:** 10.64898/2026.05.17.26353435

**Authors:** Christopher J. Gordon, Mirim Shin, Yue Leon Guo, Joanne S. Carpenter, Rebecca Robillard, Jacob J. Crouse, Sharon L. Naismith, Elizabeth M Scott, Daniel F. Hermens, Ian B. Hickie

## Abstract

Young people with major depressive disorder (MDD) exhibit altered thermoregulation, which has also been linked to vigilance and sustained attention. However, whether peripheral skin temperature is associated with cognitive vulnerability around sleep onset is unknown. We examined the relationship between the distal-proximal skin temperature gradient (DPG) and vigilance in 38 young people with MDD (20.1±3.7 years, 65.9% female) using an in-laboratory protocol spanning 4h before, to 2h after, habitual sleep time. Participants were classified into DPG_warm_ and DPG_cold_ subgroups based on being above or below median DPG before sleep onset. Linear mixed models adjusted for age and sex examined psychomotor vigilance task performance across timepoints. The DPG_warm_ subgroup (n=19) showed significantly worse performance than DPG_cold_ (n=19) across the evening for mean reaction time (RT), reciprocal reaction time, number of lapses, and fastest 10% of RT (all *p*≤0.003). Significant Group×Time interactions were observed for mean RT (F(3,90.4)=5.00, *p*=0.003) and lapses (F(3,93.6)=6.73, *p*<0.001), with DPG_warm_ participants showing progressively worse performance approaching sleep onset. At 2h post-habitual sleep onset, DPG_warm_ participants exhibited slower RT (Δ=129ms, *p*<0.001) and nearly four times more lapses (14.9 vs 4.1, *p*<0.001). Performance decrements were not accompanied by differences in melatonin timing, subjective sleepiness or mood, suggesting DPG may index cognitive vulnerability independently. Of note, younger age was associated with greater vigilance decrements. These findings demonstrate that elevated peripheral skin temperature before sleep onset is associated with reduced vigilance in young people with MDD, and may therefore have potential utility as a non-invasive thermoregulatory biomarker of cognitive vulnerability.

**Highlights:**

- Warmer peripheral skin temperature predicts vigilance decrements in young MDD
- Vigilance deficits occur without increased subjective sleepiness or mood change
- Younger age amplifies thermoregulatory-related cognitive vulnerability in MDD
- Distal-proximal skin gradient may be a non-invasive cognitive biomarker

## INTRODUCTION

Sleep, circadian rhythms, and mental health are reciprocally interlinked. Young people are especially vulnerable to delayed sleep timing due to a normative developmental shift toward later sleep-wake times (Carskadon et al., 1998; Crowley et al., 2018; Roenneberg et al., 2004) and behavioural factors, such as late evening social activities (Crowley et al., 2018), resulting in sleep-wake disturbances with implications for mental health (Crouse et al., 2021; Meyer et al., 2024; Walker et al., 2020). Young people with depressive and bipolar disorders are particularly affected by sleep and circadian rhythm disturbance (Hickie and Crouse, 2024), with evidence of considerable heterogeneity, including problems with sleep initiation and maintenance, frequent nocturnal waking, and reduced sleep quality (Asarnow and Mirchandaney, 2021). These disturbances are associated with circadian alterations, including delayed circadian phase (Carpenter et al., 2017) and internal misalignment between multiple circadian-regulated processes, not merely misalignment with the external light-dark cycle (Carpenter et al., 2025b).

Body temperature regulation is intrinsically linked to circadian rhythm function, with core temperature following a 24-hour cycle that serves as both an output of the circadian system and a signal for sleep-wake timing (Kräuchi, 2007; van Someren, 2000). The greatest rate of decline in core temperature aligns with sleep onset (Harding et al., 2020), preceded by vasodilation in distal glabrous skin regions (*e*.*g*. fingers, hands, feet) (Kräuchi et al., 2000) via arteriovenous anastomoses (Johnson and Kellogg, 2010; Rubinstein and Sessler, 1990), which increases convective heat loss and produces a distal-proximal gradient (DPG) in skin temperatures. Individuals with depressive and bipolar disorders exhibit changes in core temperature with blunted, phase-shifted, or irregular temperature curves (Avery et al., 1999; Elsenga and Van den Hoofdakker, 1988; Hasler et al., 2010; Szuba et al., 1997). Our research has also shown that young people with mood disorders exhibit distinct 24-hour skin temperature rhythms that are associated with clinical stage and mood disorder subtype (Shin et al., 2025).

Alterations in thermoregulatory function have also been associated with cognitive performance, particularly vigilance or sustained attention (Romeijn et al., 2012a). In healthy young people, normal fluctuations in skin temperature across the day are closely correlated with attention and reaction time, with better performance occurring during troughs in daytime skin temperature fluctuations (Molina et al., 2019; Romeijn and van Someren, 2011). The DPG, an indirect measure of heat flux increases and decreases indicating vasodilatory and vasoconstrictive capacity respectively, is also associated with greater vulnerability to vigilance decrements, particularly with sleep deprivation (Romeijn et al., 2012b). Controlled studies demonstrated that mild skin cooling enhances sustained task performance and wakefulness maintenance (Fronczek et al., 2008; Raymann and van Someren, 2007), while mild skin warming facilitates sleep onset and maintenance (Fronczek et al., 2008; Raymann et al., 2005), providing strong support for a causal contribution of skin temperature to vigilance regulation (Romeijn et al., 2012a).

Young people with mood disorders commonly experience significant cognitive difficulties, particularly in sustained attention and vigilance (Schumacher et al., 2024; Sommerfeldt et al., 2016), with reduced neurobehavioral performance on the psychomotor vigilance task (PVT) associated with increased depressive symptomatology (Plante et al., 2020). Our previous research of young people with affective disorders identified a cluster with longer sleep time and later wake-up time who showed significantly worse visual memory task performance (Carpenter et al., 2015).

Building on our prior work identifying skin temperature as a measure of circadian disruption in young people with mood disorders (Shin et al., 2025), we examined the relationship between DPG and vigilance in young people with major depressive disorder (MDD), around habitual sleep onset. We hypothesised that higher DPG (*i*.*e*. warmer) skin temperatures would be significantly related to worse vigilance performance compared to those with lower DPGs (*i*.*e*. cooler), skin temperatures and that these differences would become more pronounced as time since awakening increased.

## METHODS

### Participants

Data were drawn from the Youth Mental Health Follow Up Study, a transdiagnostic longitudinal study of emerging mental disorders in young people (16-40 years), conducted at the Brain and Mind Centre, University of Sydney (October 2008-November 2016) (Carpenter et al., 2017; Lee et al., 2018). Participants were recruited from primary care-based early intervention mental health services in Sydney, Australia (Scott et al., 2012) based on the presence of major mood, psychotic or developmental/behavioural syndrome. Exclusion criteria included pre-existing neurological conditions (e.g., epilepsy), medical illnesses known to affect cognitive and brain function (e.g., cancer), electroconvulsive therapy in the past three months, or current substance dependence. Additionally, those with clinically assessed impaired English language skills or intellectual disability that prevented completion of study self-ratings were excluded. The present analyses were restricted to participants with a primary diagnosis of MDD, as determined by semi-structured clinical interview conducted by a mental health professional. This study was approved by the University of Sydney Human Research Ethics Committee (2012/1631 and 2012/1628). All participants provided written informed consent.

### Procedures

Participants completed a series of baseline questionnaires assessing sleep, mood, chronotype, and social functioning. Within one month, they attended the Chronobiology and Sleep Laboratory at the Brain and Mind Centre for laboratory-based assessments. Details of the laboratory testing are described elsewhere (Carpenter et al., 2017). Briefly, participants arrived approximately 8h before habitual sleep time (HST), ascertained from baseline actigraphy recorded over 5-19 days prior to the laboratory visit. Participants swallowed an ingestible core temperature sensor and were fitted with skin temperature sensors. Thereafter, they remained in an upright seated position under controlled lighting conditions (≤30 lux) for the entire testing period, commencing at 4h before until 2h after HST. Caffeine intake was prohibited from noon on the study day, and temperature-controlled snacks and drinks were provided to minimise disruption of temperature recordings. Salivary melatonin samples were collected every 30 mins, and a psychomotor vigilance task (PVT) assessment was conducted every 2h. Participants were then permitted to sleep (at 2h after HST) and were discharged the following morning after waking at their habitual wake time (HWT).

### Clinical assessments

Prior to laboratory testing, a psychiatrist or research psychologist conducted semi-structured diagnostic interviews. Depressive symptom severity was assessed using the Hamilton Depression Rating Scale (HDRS) (Hamilton, 1960), with insomnia-related items excluded from the total score to index depressive severity independently of sleep disturbances. Functioning was assessed with the Social and Occupational Functioning Assessment Scale (SOFAS), where higher scores indicate better functioning (Goldman et al., 1992).

### Self-reported sleep assessments

Participants completed validated measures of sleep quality and chronotype before the laboratory assessment. The Pittsburgh Sleep Quality Index (PSQI) measured subjective sleep quality over the prior month (Buysse et al., 1989) and the Morningness-Eveningness Questionnaire (MEQ) assessed chronotype preferences (Horne and Ostberg, 1976).

### Laboratory assessments

#### Psychomotor Vigilance Task

Vigilance was assessed using the PVT (Dinges and Powell, 1985), administered on a validated device (PVT-192, Ambulatory Monitoring, Inc., Ardsley, NY: USA) for 10 mins every two hours from 4h before HST until 2h after HST (4 times in total). Participants responded as fast as possible to a visual stimulus; reaction time was recorded in milliseconds (msec). The inter-stimulus intervals randomly varied between 2 to 10s, and reaction times longer than 500 msec were considered lapses. We included the following PVT variables: (1) mean reaction time (RT, ms); (2) mean 1/RT (also called reciprocal reaction time (RRT, 1/s); (3) number of lapses; (4) mean of the slowest 10% of RT (ms); and (5) mean of the fastest 10% of RT (ms).

#### Core body temperature measurement

Core body temperature was monitored continuously using an ingestible temperature sensor (Equivital, Cambridge, UK) from approximately 4h before HST and continued until 3h after HWT. In these analyses, mean core body temperature was computed over the window from 4h before to 2h after HST.

#### Skin temperature measurement

Skin temperature was measured continuously throughout the study using Thermochron™ iButtons (DS1922L; Maxim Integrated Products, San Jose: CA), placed at seven body sites (finger, hand, toe, foot, abdomen, chest, forehead). Data were sampled every 60 seconds at 0.0625°C resolution. Distal skin temperature was calculated as the mean of measurements from the hand, finger, toe, and foot, while proximal temperature was derived from the forehead, chest and abdomen. The distal-proximal gradient (DPG) was computed as the difference between distal and proximal temperatures (Kräuchi et al., 2000). DPG skin temperature values were averaged over the 5 mins immediately following each PVT assessment.

#### Melatonin assessment

Salivary samples were collected every 30 mins from 6h before to 2h after HST using Salivette tubes (Sarstedt, Nümbrecht, Germany). All samples were immediately frozen and analysed later. Melatonin was assayed in duplicate using the radioimmunoassay method (Buhlmann Laboratories AG, Schnenbuch, Switzerland). This has a threshold of 0.999 pg/mL, and the inter-assay coefficient of variation is 8.2-15%. Dim light melatonin onset (DLMO) was defined as the time at which melatonin concentration first exceeded 3 pg/mL and remained above this threshold for the subsequent three consecutive samples, with linear interpolation applied between sampling intervals.

#### Subjective assessments

Subjective sleepiness and mood were assessed every 2h immediately following PVT testing using the Karolinska Sleepiness Scale (KSS) (Åkerstedt and Gillberg, 1990) and Visual Analogue Mood Scales, respectively.

### Statistical analysis

All data are presented as means ± standard deviations (SD) unless otherwise stated. Analyses proceeded in two stages. First, we visually inspected the DPG and PVT data across the -4 to +2h relative to HST. We then examined the relationship between DPG and PVT outcomes using linear regression, with DPG at -4h as the predictor variable and PVT outcomes measured at +2h as the dependent variable. Autocorrelation (Durbin-Watson) and stationarity (Augmented Dickey-Fuller) were satisfactory, and we proceeded with the next step. Due to missing PVT outcome data at +2h for some participants, regression analyses were conducted on available data per outcome (n=31-37). In a second step, participants were categorised into two DPG subgroups (DPG_warm_ and DPG_cold_) based on the median DPG values at 4h before HST. This timepoint was selected to capture inter-individual differences in thermoregulatory state prior to the rapid distal vasodilation that typically precedes sleep onset, thereby minimising confounding by circadian phase proximity and homeostatic sleep pressure. A median split (rather than continuous DPG) was used for the trajectory analyses to enable group-level visualisation, timepoint comparisons, and interpretable subgroup contrasts. Between-group comparisons for demographic and clinical characteristics were conducted using ANOVA or Kruskal-Wallis test based on normality testing, and chi-square tests for categorical variables. Linear mixed effects models were used to compare PVT outcomes between DPG_warm_ and DPG_cold_ subgroups across time points (-4, -2, 0, +2 h relative to HST), with a random intercept for participant and a first-order autoregressive residual correlation structure to account for repeated measurements. The models used all available data at each timepoint under a missing-at-random assumption; no additional imputation was performed. Four nested linear mixed models were tested to assess the robustness of findings: Model 1 (unadjusted); Model 2 adjusted for age and sex (primary model); Model 3 further adjusted for body mass index (BMI); and Model 4 adjusted for age, sex, and dim light melatonin onset (DLMO). Model 2 results are reported in the main text, with Models 1, 3, and 4 presented in Supplementary Materials. All analyses were performed using SAS 9.4 with statistical significance set at *p*<0.05.

## RESULTS

### Participants characteristics

There were 41 participants with MDD (20.1 ± 3.7 years, 65.9% female). Three participants were excluded from subsequent analyses due to missing DPG data at 4h before HST, leaving 38 participants for the analyses. Participants were categorised into DPG_warm_ (n=19) and DPG_cold_ (n=19) subgroups based on median split of DPG values. No significant differences were found between warm and cold DPG subgroups in age, BMI, sex distribution, average core body temperature, DLMO timing, or clinical measures including SOFAS, HDRS, chronotype, and PSQI scores (all *p*>0.05, Table 1).

**Table 1.**
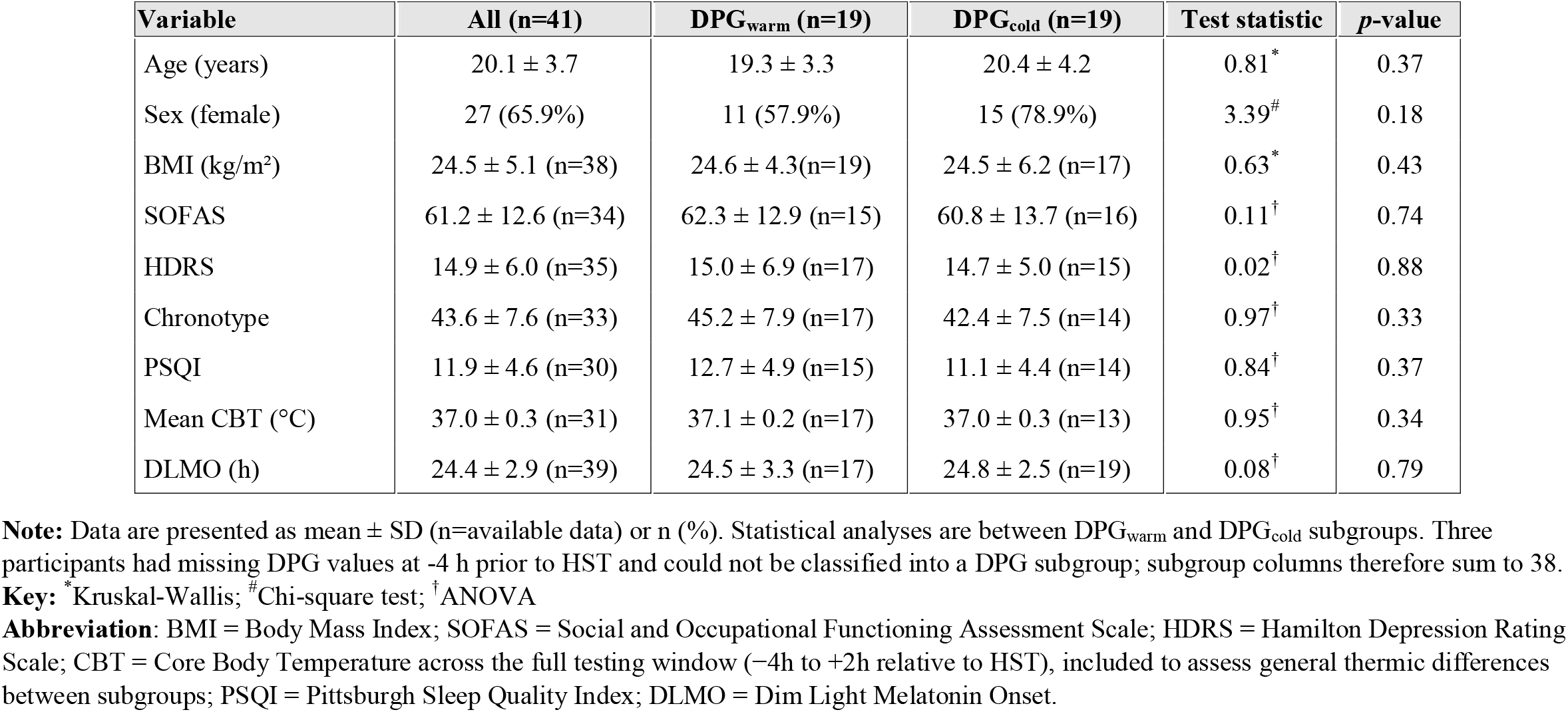
Demographic and clinical characteristics of all young people with MDD and DPG subgroups.

| Variable | All (n=41) | DPG<sub>warm</sub> (n=19) | DPG<sub>cold</sub> (n=19) | Test statistic | p-value |
| --- | --- | --- | --- | --- | --- |
| Age (years) | 20.1 ± 3.7 | 19.3 ± 3.3 | 20.4 ± 4.2 | 0.81<sup>*</sup> | 0.37 |
| Sex (female) | 27 (65.9%) | 11 (57.9%) | 15 (78.9%) | 3.39<sup>#</sup> | 0.18 |
| BMI (kg/m<sup>2</sup>) | 24.5 ± 5.1 (n=38) | 24.6 ± 4.3(n=19) | 24.5 ± 6.2 (n=17) | 0.63<sup>*</sup> | 0.43 |
| SOFAS | 61.2 ± 12.6 (n=34) | 62.3 ± 12.9 (n=15) | 60.8 ± 13.7 (n=16) | 0.11<sup>†</sup> | 0.74 |
| HDRS | 14.9 ± 6.0 (n=35) | 15.0 ± 6.9 (n=17) | 14.7 ± 5.0 (n=15) | 0.02<sup>†</sup> | 0.88 |
| Chronotype | 43.6 ± 7.6 (n=33) | 45.2 ± 7.9 (n=17) | 42.4 ± 7.5 (n=14) | 0.97<sup>†</sup> | 0.33 |
| PSQI | 11.9 ± 4.6 (n=30) | 12.7 ± 4.9 (n=15) | 11.1 ± 4.4 (n=14) | 0.84<sup>†</sup> | 0.37 |
| Mean CBT (°C) | 37.0 ± 0.3 (n=31) | 37.1 ± 0.2 (n=17) | 37.0 ± 0.3 (n=13) | 0.95<sup>†</sup> | 0.34 |
| DLMO (h) | 24.4 ± 2.9 (n=39) | 24.5 ± 3.3 (n=17) | 24.8 ± 2.5 (n=19) | 0.08<sup>†</sup> | 0.79 |

**Note:** Data are presented as mean ± SD (n=available data) or n (%). Statistical analyses are between DPG<sub>warm</sub> and DPG<sub>cold</sub> subgroups. Three participants had missing DPG values at -4 h prior to HST and could not be classified into a DPG subgroup; subgroup columns therefore sum to 38.
**Key:** <sup>\*</sup>Kruskal-Wallis; <sup>#</sup>Chi-square test; <sup>†</sup>ANOVA
**Abbreviation:** BMI = Body Mass Index; SOFAS = Social and Occupational Functioning Assessment Scale; HDRS = Hamilton Depression Rating Scale; CBT = Core Body Temperature across the full testing window (-4h to +2h relative to HST), included to assess general thermic differences between subgroups; PSQI = Pittsburgh Sleep Quality Index; DLMO = Dim Light Melatonin Onset.

### Relationship between DPG and PVT performance

Among the 38 participants, significant relationships were observed between DPG at 4h pre-HST and PVT performance at 2h post-HST (Figure 1). Higher DPG significantly predicted: (1) slower RT (β=18.52, SE=8.11, *p*=0.030, R^2^=0.15), (2) decreased RRT (β=-0.12, SE=0.045, *p*=0.010, R^2^=0.19), and (3) greater number of lapses (β=1.71, SE=0.67, *p*=0.016, R^2^=0.17). The fastest 10% RT showed marginal slowing with higher DPG (β=3.67, SE=1.97, *p*=0.072, R^2^=0.10), while the slowest 10% RT showed no significant relationship (β=202.69, SE=134.00, *p*=0.158, R^2^=0.06). No significant relationships were found between DPG at -4h and subjective sleepiness at +2h (*p*=0.304) or mood +2h (*p*=0.737).

**Figure 1.**
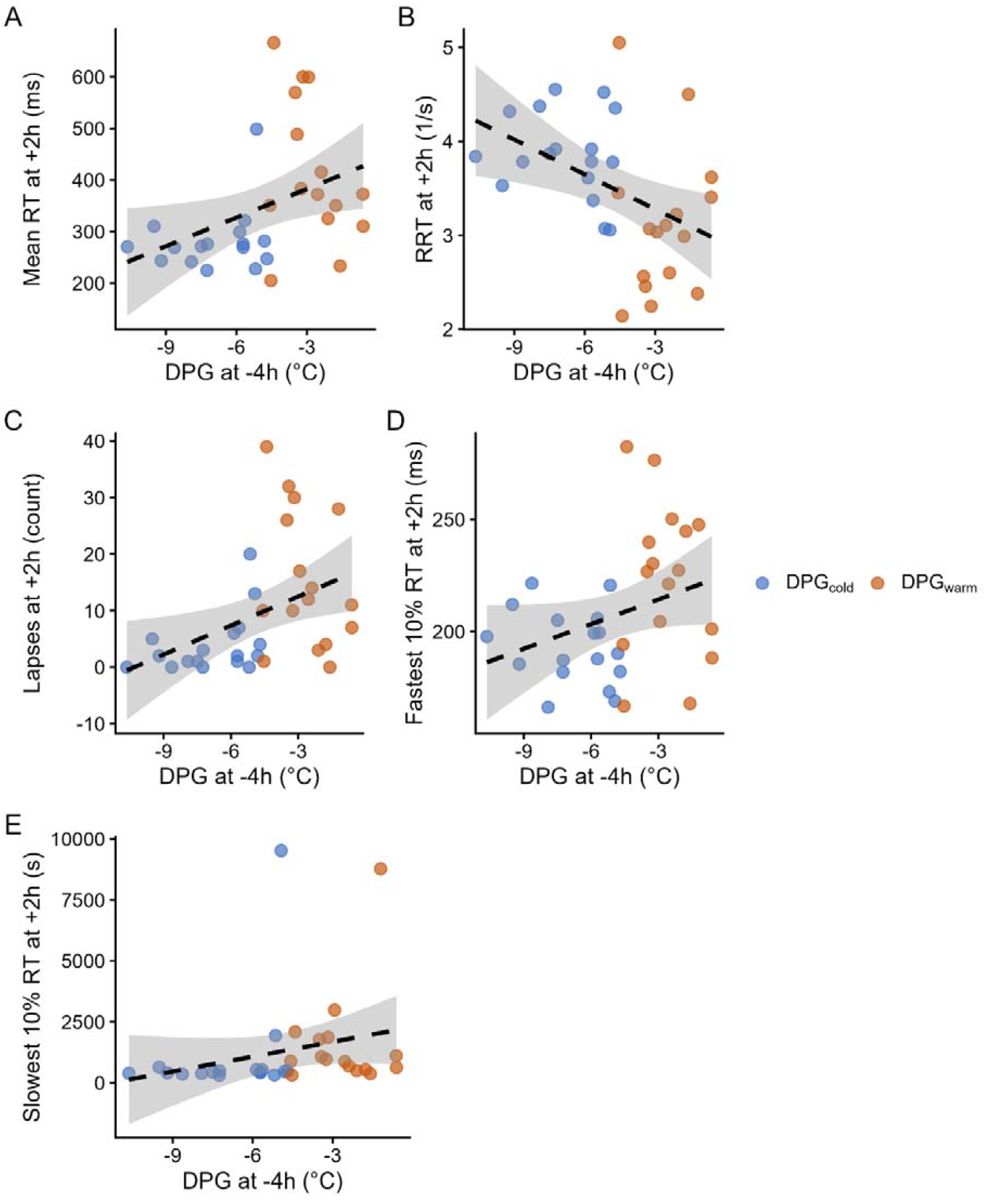
The distal-proximal gradient (DPG) of skin temperature at -4h prior to habitual bedtime and psychomotor vigilance task variables (A. reaction time, B. reciprocal reaction time, C. lapses, D. slowest and E. fastest 10% reaction times) at 2h after habitual bedtime in young people with depressive disorders (n=31-38, varying by outcome due to missing data).

### Comparison between DPG_warm_ and DPG_cold_ subgroups

Linear mixed effects models examined vigilance performance between DPG subgroups across the four time points (-4, -2, 0, +2h relative to HST). Results from the primary model (Model 2: adjusted for age and sex) are reported below; unadjusted and sensitivity analyses (Models 1, 3, 4) are presented in Supplementary Tables.

Significant main effects of group were observed for mean RT (F(1,33.5)=15.84, *p*=0.0004), RRT (F(1,37.5)=10.76, *p*=0.002), lapses (F(1,33.4)=10.26, *p*=0.003), and fastest 10% RT (F(1,38.8)=11.23, *p*=0.002), with the DPG_warm_ subgroup consistently showing poorer vigilance performance across these measures. No significant group effect was observed for slowest 10% RT (F(1,35.2)=0.33, *p*=0.571). Significant main effects of time were observed for all PVT measures (all *p*≤0.004). No significant group effects were found for subjective sleepiness (F(1,40.3)=0.85, *p*=0.363) or mood (F(1,35.3)=1.00, *p*=0.324).

Significant Group × Time interactions were observed for mean RT (F(3,90.4)=5.00, *p*=0.003) and lapses (F(3,93.6)=6.73, *p*<0.001), indicating that group differences in these measures increased progressively across the evening (Figure 2). The interactions for RRT (F(3,100.5)=1.47, *p*=0.228), fastest 10% RT (F(3,100.1)=2.01, *p*=0.117), and slowest 10% RT (F(3,103.5)=0.26, *p*=0.852) were not significant, suggesting that group differences in these measures remained relatively stable across timepoints. No significant interactions were observed for KSS (*p*=0.602) or mood (*p*=0.904).

**Figure 2.**
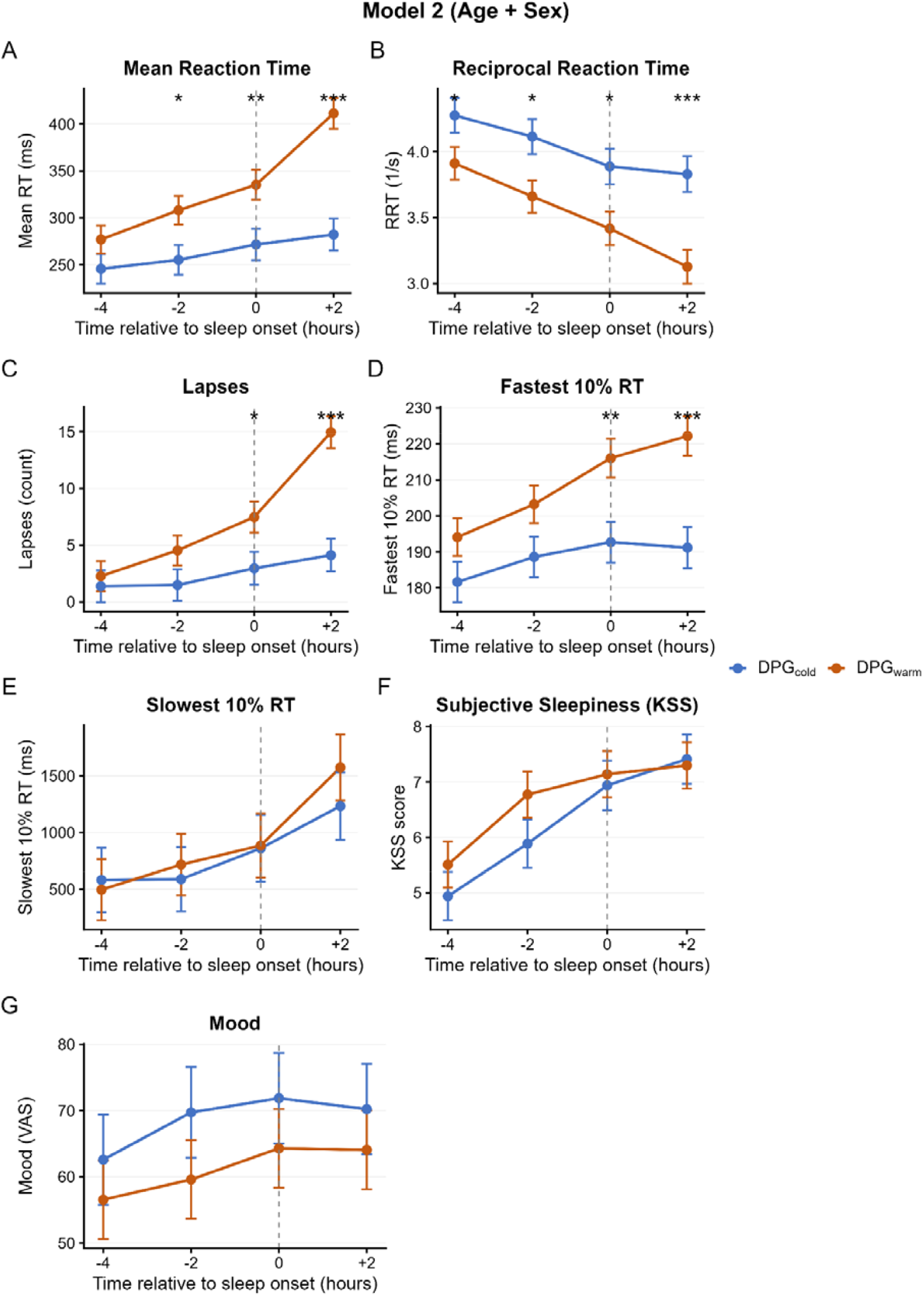
Psychomotor vigilance task outcomes (reaction time, reciprocal reaction time, lapses, fastest and slowest 10%), subjective sleepiness and mood (VAS) before and after habitual sleep time in MDD DPG_warm_ (n=19) and DPG_cold_ subgroups (n=19). Data are model-estimated means ± standard error. Vertical dashed line indicates habitual sleep time (time 0). Asterisks above panels denote significant post-hoc group differences at each timepoint: * *p*<0.05; ** *p*<0.01; *** *p*<0.001.

For outcomes with significant Group × Time interactions, Tukey-adjusted post-hoc comparisons examined group differences at each timepoint (Figure 2). For mean RT, the DPG_warm_ subgroup showed significantly slower responses than DPG_cold_ at -2h (Δ=52.9 ms, *p*=0.021), 0h (Δ=63.6 ms, *p*=0.009), and +2h (Δ=129.0 ms, *p*<0.001), but not at -4h (*p*=0.158). The magnitude of group differences increased progressively, with the largest difference observed at 2h post-HST. For lapses, significant group differences emerged at 0h (Δ=4.5 lapses, *p*=0.028) and +2h (Δ=10.8 lapses, *p*<0.0001), but not at earlier timepoints (-4h: *p*=0.640; -2h: *p*=0.116). At 2h post-HST, the DPG_warm_ subgroup exhibited nearly four times as many lapses as the DPG_cold_ subgroup (14.9 ± 1.4 vs 4.1 ± 1.4 lapses; *p*<0.001).

Age was a significant covariate for mean RT (*p*=0.010), RRT (*p*=0.013), lapses (*p*=0.022), indicating that younger participants exhibited greater vigilance decrements. Age showed a marginal trend for slowest 10% RT (F(1,34.4)=4.11, *p*=0.050). Sex was a significant covariate for fastest 10% RT (F(1,38.6)=4.35, *p*=0.044) but not for any other outcome (all *p*>0.05). Results were consistent across sensitivity analyses additionally adjusting for BMI (Model 3) or DLMO (Model 4), with significant Group × Time interactions for mean RT and lapses observed across all four models (see Supplementary Tables).

## DISCUSSION

This is the first study to examine peripheral thermoregulation and vigilance in young people with mood disorders. Young people with MDD who had warmer pre-sleep peripheral skin temperatures showed substantially reduced vigilance—slower mean and fastest RTs, reduced RRT, and more lapses—than those with cooler temperatures. Critically, for mean RT and lapses, this decrement became progressively more pronounced as bedtime approached. We showed that DPG skin temperature four hours before habitual sleep time predicted slower and more error-prone vigilance performance two hours after habitual bedtime. Notably, the vigilance performance decrements in the DPG_warm_ subgroup were divergent by group differences in subjective sleepiness or mood, suggesting that altered thermoregulatory physiology may serve as an index of cognitive vulnerability in young people with MDD around sleep onset.

These findings extend previous research in healthy populations, demonstrating associations between higher skin temperatures and reduced vigilance (Molina et al., 2019; Romeijn and van Someren, 2011). In healthy individuals, the nocturnal increase in DPG mediates sleep onset through pronounced distal convective heat loss, facilitating the maximal decline in core temperature (Kräuchi et al., 2000). In healthy people, lower vigilance approaching sleep onset is itself a normative, adaptive feature reflecting the homeostatic and circadian coupling between thermoregulation and arousal regulation (Kräuchi, 2007). In our study, an early and elevated DPG signalled a decline in the ability to maintain vigilant attention. The significant interactions for PVT performance show that for the DPG_warm_ subgroup, the biological drive for sleep or the circadian-mediated decline in alertness may be more strongly coupled with distal thermoregulatory-mediated vasodilation. Furthermore, our observation that younger participants showed greater vigilance decrements aligned with the known developmental sensitivity of young adults to circadian and sleep-wake disturbances (Carskadon et al., 1998; Crowley et al., 2018).

The heterogeneity in vigilance performance observed here aligns with our previous identification of distinct sleep-wake and physiological subgroups in young people with mood disorders. In a partially overlapping sample, Carpenter et al. (2015) identified a cluster characterised by longer sleep duration and later wake times whose members showed worse visual memory performance relative to a disrupted sleep cluster. The present divergence between DPG_warm_ and DPG_cold_ subgroups mirrors this pattern. While that earlier work focused on sleep macroarchitecture, the current findings suggest that the identified cognitive deficits may relate to thermoregulatory differences in heat dissipation before sleep onset. The reduced vigilance observed in the DPG_warm_ subgroup also resonates with the position of cognitive performance as a downstream output of circadian dysregulation within the broader ‘circadian depression’ framework (Carpenter et al., 2021). However, given that the DPG subgroups did not differ on DLMO, chronotype, or subjective sleep disturbance—core markers of that phenotype—whether thermoregulatory-mediated cognitive vulnerability represents a distinct dimension of circadian pathophysiology or overlaps with circadian depression as currently defined remains to be explored further. Taken together, DPG may provide a physiological mechanism by which some individuals with MDD experience cognitive fog while others remain relatively unaffected, despite similar clinical presentations (Sommerfeldt et al., 2016).

Whilst there were no differences in DLMO between the DPG subgroups, the increased distal vasodilation observed in the DPG_warm_ subgroup may reflect a decoupling between sleep-wake homeostasis and circadian alignment. Importantly, the DPG subgroups were defined at 4h before HST, in advance of the homeostatic pre-sleep distal vasodilation in healthy adults; elevated DPG at this timepoint is therefore unlikely to reflect a more advanced stage of normal pre-sleep physiology, but may instead indicate an earlier-than-expected ramp consistent with internal circadian misalignment (Carpenter et al., 2025a). Whilst it has been postulated that melatonin release drives the increase in DPG prior to sleep onset (Kräuchi K et al., 1997), this single circadian marker may not fully capture circadian contributions to the vigilance effects observed in the current study. Our previous work has shown some young people with mood disorders could be grouped as a delayed sleep-wake subtype with corresponding delayed melatonin onset and core temperature nadir (Carpenter et al., 2017). This is further reinforced by our finding that 24-hour skin temperature rhythms are sensitive indicators of circadian disruption in young people with mood disorders (Shin et al., 2025). This circadian-sleep-wake misalignment may negatively affect attentional networks that are sensitive to sleep loss and circadian timing (Tomasi et al., 2009). Extending this, Carpenter et al. (2025) demonstrated that internal circadian misalignment in young people with mood disorders is driven largely by earlier relative core temperature rhythms rather than delayed melatonin—consistent with the present observation that thermoregulatory patterning, rather than DLMO, tracks vigilance vulnerability. DPG may therefore serve as a surrogate of circadian-thermoregulatory integrity, offering a non-invasive marker of cognitive risk beyond what melatonin indices may provide. Whether this reflects a general trait-level vulnerability or a circadian-phase-specific effect confined to the pre-sleep window remains to be determined. Notably, the DPG_cold_ subgroup may represent a physiologically typical pre-sleep thermoregulatory state rather than active cognitive resilience—a distinction that cannot be resolved without a healthy control comparison. Equally, we cannot rule out the possibility that the DPG_cold_ subgroup reflects subtle alterations in sleep-wake regulation or thermoregulatory function that contribute to sleep initiation difficulties, although the absence of group differences in subjective sleepiness and mood suggests any such effects are not strongly mirrored at the subjective level.

So how does an elevated DPG associated with sleep timing result in increased peripheral thermoafferents that negatively affect vigilant attention networks? Research has shown that thermosensitive neurons in preoptic anterior hypothalamus (POAH) and peripheral thermoafferents regulate mean body temperature while influencing sleep-wake cycles (Gilbert et al., 2004; Nakamura and Morrison, 2008; Tan et al., 2016). Animal studies show that increasing peripheral warming increases thermoafferent signalling via the lateral parabrachial nucleus to the median preoptic nucleus and medial preoptic area, activating warm-thermosensitive POAH neurons that initiate sleep onset and maintain non-rapid eye movement sleep (Alam et al., 1995; Harding et al., 2018). These neurons project to the GABAergic ventrolateral preoptic (VLPO) neurons, which promote sleep by inhibiting the ascending arousal system (Harding et al., 2019). Such thermoregulatory signalling also inhibits higher-order attentional neuronal networks. As sleep propensity increases, these bottom-up processes dominate top-down attentional control due to sleep loss (Van Dongen et al., 2011). However, the exact neurobiological underpinnings of how peripheral thermoafferents mediate decrements in vigilance remains to be fully elucidated.

The differentiation of the vigilance-sensitive DPG subgroups suggests that peripheral thermoregulatory signatures may reflect a trait-like vulnerability in neurobehavioral performance, consistent with the inter-individual differences in sleep loss sensitivity previously identified (Van Dongen et al., 2004; Van Dongen et al., 2012). Although this framework was developed in the context of healthy individuals undergoing experimental sleep deprivation while attempting to maintain wakefulness, rather than in the natural pre-sleep window, the underlying concept of stable inter-individual variability in neurobehavioral response to sleep-wake pressure may extend to the present setting. The DPG_cold_ subgroup may represent a resilient subtype who maintain relatively stable vigilance with increasing sleep loss. Conversely, the DPG_warm_ subgroup typifies the vulnerable subtype, characterised by a progressive and significant reduction in sustained attention due to homeostatic sleep pressure. This divergence positions the DPG as a sensitive physiological marker of inter-individual variability and a potential biomarker of cognitive vulnerability in young people with MDD.

There are several limitations in this study. First, the design is observational and restricted to the evening pre-sleep window; causality cannot be established, and we cannot determine whether warmer DPG drives vigilance decrements or whether both reflect a common underlying process such as arousal-system dysregulation. Second, the sample size was modest, and the median split approach, while clinically interpretable, reduces statistical information relative to continuous modelling; future analyses should prioritise continuous DPG measures and explore non-linear or time-varying effects. Third, the controlled laboratory setting—fixed posture, lighting and repeated PVT testing—increases internal validity but may not reflect real-world evening contexts. Fourth, MDD is a diagnostically heterogeneous condition, and differences in illness stage and psychotropic medication use may have moderated the DPG-vigilance relationship in ways the current sample could not fully examine. Finally, without a healthy control comparison in the present analyses, we cannot determine whether the observed DPG-vigilance coupling is specific to MDD or represents a general feature of approaching bedtime.

Future studies should investigate the mechanistic underpinnings of the skin temperature-vigilance relationship by combining peripheral and core temperature with autonomic measures (*e*.*g*. heart rate variability) and neurophysiological indices of arousal, including electroencephalographic markers of objective sleepiness where waking EEG data quality permits, to test shared pathways linking thermal afference and vigilance regulation (Romeijn et al., 2012b). Circadian modelling to quantify 24-hour rhythms and internal phase relationships among temperature, melatonin, and sleep would provide insights into how these interact especially in the different DPG subgroups. An experimental manipulation of distal skin temperature (*e*.*g*., cooling versus thermoneutral) during the pre-sleep window, whilst measuring PVT outcomes and next-day functioning, would allow examination of whether this is a state effect. Notably, not all circadian interventions improve vigilance. Bright light inhibits melatonin release but did not improve PVT performance in a recent study (Lazar et al., 2025), highlighting the need for mechanistically-targeted thermal approaches.

In conclusion, peripheral thermoregulation in young people with MDD is associated with vigilance vulnerability around sleep onset. While elevated DPG typically reflects greater sleep propensity, the DPG_warm_ subgroup may experience greater cognitive vulnerability at this transition, potentially via thermoafferent disruption of attentional pathways. Further work in real-world settings and through thermal manipulation is needed to clarify causality.

## Data Availability

All data produced in the present study are available upon reasonable request to the authors

## Acknowledgements

We would like to express our gratitude to the participants whose time and cooperation were crucial to this study. Furthermore, we wish to acknowledge the research staff for their support.

